# Safety and immunogenicity of a heterologous boost with a recombinant vaccine, NVSI-06-07, in the inactivated vaccine recipients from UAE: a phase 2 randomised, double-blinded, controlled clinical trial

**DOI:** 10.1101/2021.12.29.21268499

**Authors:** Nawal AlKaabi, Yun Kai Yang, Jing Zhang, Ke Xu, Yu Liang, Yun Kang, Ji Guo Su, Tian Yang, Salah Hussein, Mohamed Saif ElDein, Shuai Shao, Sen Sen Yang, Wenwen Lei, Xue Jun Gao, Zhiwei Jiang, Hui Wang, Meng Li, Hanadi Mekki Mekki, Walid Zaher, Sally Mahmoud, Xue Zhang, Chang Qu, Dan Ying Liu, Jing Zhang, Mengjie Yang, Islam ElTantawy, Peng Xiao, Zhao Nian Wang, Jin Liang Yin, Xiao Yan Mao, Jin Zhang, Ning Liu, Fu Jie Shen, Liang Qu, Yun Tao Zhang, Xiao Ming Yang, Guizhen Wu, Qi Ming Li

## Abstract

**Background:** The increased coronavirus disease 2019 (COVID-19) breakthrough cases pose the need of booster vaccinations. In this study, we reported the safety and immunogenicity of a heterologous boost with a recombinant COVID-19 vaccine (CHO cells), named NVSI-06-07, as a third dose in participants who have previously received two doses of the inactivated vaccine (BBIBP-CorV) at pre-specified time intervals. Using homologous boost with BBIBP-CorV as control, the safety and immunogenicity of the heterologous boost with NVSI-06-07 against various SARS-CoV-2 strains, including Omicron, were characterized.

**Methods:** This study is a single-center, randomised, double-blinded, controlled phase 2 trial for heterologous boost of NVSI-06-07 in BBIBP-CorV recipients from the United Arab Emirates (UAE). Healthy adults (aged ≥18 years) were enrolled and grouped by the specified prior vaccination interval of BBIBP-CorV, i.e., 1-3 months, 4-6 months or ≥6 months, respectively, with 600 individuals per group. For each group, participants were randomly assigned at 1:1 ratio to receive either a heterologous boost of NVSI-06-07 or a homologous booster dose of BBIBP-CorV. The primary outcome was to comparatively assess the immunogenicity between heterologous and homologous boosts at 14 and 28 days post-boosting immunization, by evaluation of the geometric mean titers (GMTs) of IgG and neutralizing antibodies as well as the corresponding seroconversion rate (≥4-fold rise in antibody titers). The secondary outcomes were the safety profile of the boosting strategies within 30 days post vaccination. The exploratory outcome was the immune efficacy against Omicron and other variants of concern (VOCs) of SARS-CoV-2. This trial is registered with ClinicalTrials.gov, NCT05033847.

**Findings:** A total of 1800 individuals who have received two doses of BBIBP-CorV were enrolled, of which 899 participants received a heterologous boost of NVSI-06-07 and 901 received a homologous boost for comparison. No vaccine-related serious adverse event (SAE) and no adverse events of special interest (AESI) were reported. 184 (20·47%) participants in the heterologous boost groups and 177 (19·64%) in the homologous boost groups reported at least one adverse reaction within 30 days. Most of the local and systemic adverse reactions reported were grades 1 (mild) or 2 (moderate), and there was no significant difference in the overall safety between heterologous and homologous boosts. Immunogenicity assays showed that the seroconversion rates in neutralizing antibodies against prototype SARS-CoV-2 elicited by heterologous boost were 89·96% - 97·52% on day 28 post-boosting vaccination, which was much higher than what was induced by homologous boost (36·80% - 81·75%). Similarly, in heterologous NVSI-06-07 booster groups, the neutralizing geometric mean titers (GMTs) against the prototype strain increased by 21·01 - 63·85 folds from baseline to 28 days post-boosting vaccination, whereas only 4·20 - 16·78 folds of increases were observed in homologous BBIBP-CorV booster group. For Omicron variant, the neutralizing antibody GMT elicited by the homologous boost of BBIBP-CorV was 37·91 (95%CI, 30·35-47·35), however, a significantly higher level of neutralizing antibodies with GMT 292·53 (95%CI, 222·81-384·07) was induced by the heterologous boost of NVSI-06-07, suggesting that it may serve as an effective boosting strategy combating the pandemic of Omicron. The similar results were obtained for other VOCs, including Alpha, Beta and Delta, in which the neutralizing response elicited by the heterologous boost was also significantly greater than that of the homologous boost. In the participants primed with BBIBP-CorV over 6 months, the largest increase in the neutralizing GMTs was obtained both in the heterologous and homologous boost groups, and thus the booster vaccination with over 6 months intervals was optimal.

**Interpretation:** Our findings indicated that the heterologous boost with NVSI-06-07 was safe, well-tolerated and immunogenic in adults primed with a full regimen of BBIBP-CorV. Compared to homologous boost with a third dose of BBIBP-CorV, incremental increases in immune responses were achieved by the heterologous boost with NVSI-06-07 against SARS-CoV-2 prototype strain, Omicron variant, and other VOCs. The heterologous BBIBP-CorV/NVSI-06-07 prime-boosting vaccination may be valuable in preventing the pandemic of Omicron. The optimal booster strategy was the heterologous boost with NVSI-06-07 over 6 months after a priming with two doses of BBIBP-CorV.

**Research in context:** *Evidence before this study:* We searched PubMed for clinical trials or prospective/cohort studies involving heterologous booster vaccination in non-immunocompromised population published up to Dec 25, 2021, using the term “(COVID) AND (vaccin*) AND (clinical trial OR cohort OR prospective) AND (heterologous) AND (booster OR prime-boost OR third dose)” with no language restrictions. Nine studies of heterologous prime-boost vaccinations with adenovirus-vector vaccines (ChAdOx1 nCov-19, Oxford-AstraZeneca, Ad26.COV2.S, Janssen) and mRNA vaccines (BNT162b2, Pfizer-BioNtech; mRNA1273, Moderna) were identified. The adenovirus-vector and mRNA heterologous prime-boost vaccination was found to be well tolerated and immunogenic. In individuals primed with adenovirus-vector vaccine, mRNA booster vaccination led to greater immune response than homologous boost. However, varied results were obtained on whether heterologous boost was immunogenically superior to the homologous mRNA prime-boost vaccination. Besides that, A preprint trial in population previously immunized with inactivated vaccines (CoronaVac, Sinovac Biotech) showed that the heterologous boost with adenovirus-vector vaccine (Convidecia, CanSino Biologicals) was safe and induced higher level of live-virus neutralizing antibodies than by the homogeneous boost. A pilot study reported that boosting with BNT162b2 in individuals primed with two doses of inactivated vaccines (BBIBP-CorV) was significantly more immunogenic than homologous vaccination with two-dose of BNT162b2. In addition, a preprint paper demonstrated that heterologous boost of ZF2001, a recombinant protein subunit vaccine, after CoronaVac or BBIBP-CorV vaccination potently improved the immunogenicity. But only a small size of samples was tested in this study and the live-virus neutralization was not detected. Till now, it is still lacking a formal clinical trial to evaluate the immunogenicity and safety of the heterologous prime-boost vaccination with an inactivated vaccine followed by a recombinant protein subunit-based vaccine.

*Added value of this study:* To our knowledge, this is the first reported result of a large-scale randomised, controlled clinical trial of heterologous prime-boost vaccination with an inactivated vaccine followed by a recombinant protein subunit vaccine. This trial demonstrated that the heterologous prime-booster vaccination with BBIBP-CorV/NVSI-06-07 is safe and immunogenic. Its immunoreactivity is similar to that of homologous vaccination with BBIBP-CorV. Compared to homologous boost, heterologous boost with NVSI-06-07 in BBIBP-CorV recipients elicited significantly higher immunogenicity not only against the SARS-CoV-2 prototype strain but also against Omicron and other variants of concern (VOCs).

*Implications of all the available evidence:* Booster vaccination is considered an effective strategy to improve the protection efficacy of COVID-19 vaccines and control the epidemic waves of SARS-CoV-2. Data from our trial suggested that the booster vaccination of NVSI-06-07 in BBIBP-CorV recipients significantly improved the immune responses against various SARS-CoV-2 strains, including Omicron. Due to no Omicron-specific vaccine available currently, the BBIBP-CorV/NVSI-06-07 heterologous prime-boost might serve as an effective strategy combating Omicron variant. Besides that, BBIBP-CorV has been widely inoculated in population, and thus further boosting vaccination with NVSI-06-07 is valuable in preventing the COVID-19 pandemic. But further studies are needed to assess the long-term protection of BBIBP-CorV/NVSI-06-07 prime-booster vaccination.

## INTRODUCTION

The epidemic of coronavirus disease 2019 (COVID-19), caused by severe acute respiratory syndrome coronavirus 2 (SARS-CoV-2), has stimulated global efforts to develop safe and effective vaccines against the rapid spread of the virus. So far, great progresses have been achieved, and a total of ten vaccines have been approved by the world health organization (WHO) for emergency use.^1^ These COVID-19 vaccines have been used in large-scale populations and shown to offer effective protections against severe disease, hospitalization and death.^2^ However, due to the waning of neutralization titer over time in vaccinated individuals and emergence of SARS-CoV-2 variants such as Omicron and Delta, breakthrough infection cases continuously increase,^3,4^ which raises the urgent need of new strategies to cope with this tendency.

Booster vaccination may be an effective way to improve waning immunity and broaden protective immune responses against SARS-CoV-2. The clinical trials in adults who have received the two-dose primary vaccination series with mRNA-1273 or BNT162b2 vaccines showed that a booster injection of the same vaccine, six to eight months later, yielded 3·8- to 7-fold higher neutralizing antibody titers against the wild-type virus compared to the peak value after the primary series.^5-7^ Besides the homologous boosting, heterologous booster strategy has also attracted great concerns, and several clinical trials and cohort studies have shown that the heterologous prime-booster vaccination was immunologically superior than homologous counterparts.^8-12^ However, the immunogenicity of the heterologous prime-booster vaccination with the inactivated vaccine and the recombinant subunit vaccine was evaluated recently only in a small size of samples, and the neutralizing antibodies against live SARS-CoV-2 were not detected in the study.^11,12^ Till now, it is still lacking a large-scale clinical trial to evaluate the immunogenicity of heterologous boost with the recombinant subunit vaccine in individuals primed with the inactivated vaccine. Besides that, the neutralization ability of heterologous booster vaccination against the newly epidemic Omicron variant also needs to be tested.

Based on structural and computational analysis of spike receptor-binding domain (RBD) of SARS-CoV-2, we have designed a recombinant COVID-19 vaccine (CHO cells), named NVSI-06-07, that uses a trimeric form of RBD as the antigen. The phase 1/2 clinical trial conducted in China has demonstrated high safety and strong immunogenicity of this vaccine. We sought to know whether the use of NVSI-06-07 as a heterologous booster vaccination can effectively improve the immune responses in the inactivated vaccine recipients. Here, we report the safety and immunogenicity of heterologous booster vaccination with NVSI-06-07 at pre-specified time intervals in individuals who have previously received two doses of the inactivated vaccine BBIBP-CorV, which were then compared to those of homologous boosting strategy with a third dose of BBIBP-CorV. Moreover, as an exploratory study, the live-virus neutralization activities of the vaccinated sera were also evaluated against Omicron and other SARS-CoV-2 variants of concern (VOCs).

## METHODS

### Trial Design and Participants

This trial was designed as a phase 2, randomised, double-blinded, controlled trial conducted at a single clinical site in UAE. Eligible participants were healthy adults, aged ≥18 years old, who had previously received a full series (two doses) of BBIBP-CorV, a COVID-19 inactivated vaccine. Three groups of participants, receiving their second dose of BBIBP-CorV 1-3 months, 4-6 months or at least 6 months ago, respectively, were enrolled with 600 individuals per group. Female volunteers were not pregnant or breastfeeding, and appropriate contraceptive measures had been taken within 2 weeks before enrollment. Participants were screened for health status by inquiry and physical examination, prior to enrollment. Volunteers who had a history of SARS-CoV-2, SARS or MERS infection, or received any COVID-19 vaccine other than BBIBP-CorV were excluded. Other exclusion criteria can be found on ClinicalTrials.gov (NCT05033847).

The trial protocol was reviewed and approved by Abu Dhabi Health Research and Technology Ethics Committee. The trial was performed in accordance with Good Clinical Practice (GCP), Declaration of Helsinki (with amendments) as well as the local legal and regulatory requirements, and trial safety was overseen by an independent safety monitoring committee. Written informed consent was provided for all participants prior to inclusion into the trial.

### Randomisation and masking

Eligible participants were grouped into three groups, i.e., 1-3 months group, 4-6 months group and ≥6 months group, according to the time interval between their study day 0 and prior vaccination date of the second dose of BBIBP-CorV. For each group, a random table was generated by block randomization method using SAS software (version 9·4). Each enrolled participant was randomly assigned to a code and to receive either a heterologous booster dose of NVSI-06-07 or a homologous booster dose of BBIBP-CorV. The trial is double-blind to avoid introducing bias by having randomization and masking process handled by independent personnel from trial operation. Participants, investigators and other staffs remained blinded to individual treatment assignment during the trial.

### Procedures

NVSI-06-07, a recombinant COVID-19 vaccine (CHO cells), encoding a trimeric form of RBD, was developed by the National Vaccine and Serum Institute (NVSI) and manufactured by Lanzhou Institute of Biological Products Co., Ltd. (LIBP) in accordance with good manufacturing practice (GMP). This vaccine is in the liquid form of 0·5 ml per dose, containing 20μg antigen and 0·3mg aluminum hydroxide adjuvant. The inactivated vaccine BBIBP-CorV, used as a control in this trial, was provide by Beijing Institute of Biological Products Co., Ltd. (BIBP), which has been approved by WHO for emergency use and applied in large populations. All vaccines were stored at 2°C-8°C prior to use.

After screening, eligible participants received the booster inoculation intramuscularly with NVSI-06-07 or BBIBP-CorV, followed by clinical observation at the study site for no less than 30 minutes. Within the subsequent 7 days after booster vaccination, local and systemic adverse events (AEs) were self-reported daily by participants using standardized diary cards and verified by investigators. From day 8 to day 30 post-vaccination, unsolicited AEs were recorded by participants in contact cards. Serious adverse events (SAEs) and adverse events of special interest (AESIs) were monitored up to 6 months after vaccination. The grade of AEs was assessed according to the relevant guidance of China National Medical Products Administration (NMPA).

The immunogenicity was assessed by RBD-specific binding antibody responses (IgG) and neutralizing antibody activities against live SARS-CoV-2 virus. Blood samples were collected from the participants before booster vaccination, and on days 14 and 28 after boost. IgG level specific to prototype RBD was measured using ELISA kits (purchased from Bioscience (Chongqing) Biotechnology Co. Ltd.). Neutralizing antibody titer was detected using live-virus neutralization assay as described in our previous studies.^13^ In order to evaluate cross-neutralizing activities, both prototype SARS-CoV-2 live virus and several VOCs, including Omicron, Alpha, Beta and Delta strains, were used in the neutralization assay. The corresponding seroconversion rates, defined as ≥4-fold rise in IgG or neutralizing titers were determined based on the detected IgG or neutralizing antibody titers.

### Outcomes

The primary outcome was the comparative assessment of immunogenicity between heterologous and homologous booster vaccinations on days 14 and 28 post-boosting. The secondary outcomes were safety profile within 30 days pos-boosting. The exploratory outcome was the immunity against Omicron and other VOCs. Safety was assessed by the occurrence of all SAEs and AESIs, and the occurrence of the solicited or unsolicited adverse reactions within 30 days after vaccination. The occurrence and severity of adverse reactions were compared between heterologous NVSI-06-07 booster groups and the homologous BBIBP-CorV booster groups. The immunogenicity was evaluated by geometric mean titers (GMTs) and the seroconversion rate (≥4-fold rise in titers) of RBD-binding antibody IgG and live-virus neutralizing antibodies on 14 and 28 days after booster vaccination. The comparisons of the immunogenicity between the heterologous and homologous booster groups were also carried out. The immunity against Omicron and other VOCs evaluated was determined using live-virus neutralizing antibody GMTs.

### Statistical analysis

Assuming that a 4-fold rise in neutralizing antibody titers for both heterogeneous and homologous booster groups reached at 85% and the non-inferiority threshold was set to -10%, the sample size was determined to be 208 using Miettinen & Nurminen method to achieve 80% power at one-sided significance level of 2·5%. Assuming that the neutralizing antibody GMTs between heterologous and homologous boosting groups are comparable, with the standard deviation (SD) of GMT after log10 transformation to be 0·7, and the non-inferiority threshold was set to 2/3, 250 participants per group was needed to achieve 80% power at one-sided significance level of 2·5%. Considering the above estimations and 15%∼20% drop-out rate, 600 participants were enrolled into each of the three boosting groups (1-3 months, 4-6 months and ≥6 months). Half participants of each group were assigned to heterologous booster and the other half were assigned to homologous booster. Thus, a total of 1800 individuals (900 in heterologous groups and 900 in homogeneous groups) participated the trial.

Baseline characteristics were evaluated in the full analysis set (FAS). Continuous variables were analyzed using Student’s t-test and categorical variables were analyzed with Chi-square test. Safety analysis was performed on the safety set (SS), and immunogenicity analysis was carried out on Per-protocol set (PPS). RBD-specific IgG levels and the neutralizing antibody activities against the live virus were presented by GMTs with 95% confidence intervals (CIs) calculated using Clopper-Pearson method. Additionally, based on pre-booster and post-booster titers, 4-fold increase in antibody titers were calculated. Cochran-Mantel-Haenszel (CMH) method considering stratification factors was used to compare the proportion differences between heterologous booster groups and homologous booster groups, and among groups with different boosting intervals. RBD-specific IgG and the neutralizing antibody titers between the heterologous and homologous booster groups were compared after logarithmic conversion. For safety analysis, the number and proportion of participants reporting at least one adverse reaction post-vaccination were analyzed and differences between groups were compared using Fisher’s exact test. All statistical analyses were carried out using SAS software (version 9·4). All statistical tests were two-sided, and the statistical significance level was *P*<0·05.

### Role of the funding source

The funder of the study had no role in trial design, data collection, data analysis, data interpretation, or writing of the paper. All authors had full access to study data and the corresponding authors had final responsibility for the decision to submit for publication.

## RESULTS

Healthy adults aged ≥18 years who received a full regimen (two doses) of BBIBP-CorV 1-3 months, 4-6 months and ≥6 months ago, respectively, were recruited as shown in Figure 1. For these three groups with different boosting intervals, a total of 1800 participants were enrolled with 600 of each group at one clinical site in UAE. For each group, participants were randomly assigned to receive either a heterologous booster vaccination with NVSI-06-07 or a homologous booster with a third dose of BBIBP-CorV (Figure 1). Demographic characteristics were similar between the heterologous and homologous boosting groups. The participants in the two groups exhibited balanced distributions in age, sex, height and body weight (Table 1). The nationality of participants was provided in Appendix Table A1. All the 1800 participants receiving booster vaccination were included in SS for safety analysis. A total of 1672 participants completed the follow-up visit on day 14, and these individuals were included in per-protocol set 1 (PPS1) for day 14 immunogenicity analysis. A total of 1496 participants completed day 28 visit, which were included in per-protocol set 2 (PPS2) for day 28 immunogenicity analysis (Figure 1).

**Table 1:** Demographic characteristics of the participants (FAS)

|  | 1-3 months |  |  | 4-6 months |  |  | ≥6 months |  |  |
| --- | --- | --- | --- | --- | --- | --- | --- | --- | --- |
|  | NVSI-06-07<br>(N=301) | BBIBP-CorV<br>(N=299) | P value | NVSI-06-07<br>(N=300) | BBIBP-CorV<br>(N=300) | P value | NVSI-06-07<br>(N=298) | BBIBP-CorV<br>(N=302) | P value |
| Age (years) |  |  |  |  |  |  |  |  |  |
| Mean (SD) | 33·30 (8·88) | 33·43 (9·38) | 0·8642 | 34·10 (7·89) | 34·53 (8·59) | 0·5159 | 35·48 (9·53) | 36·12 (9·27) | 0·4036 |
| Median | 31·88 | 32·50 |  | 33·37 | 33·72 |  | 34·08 | 34·72 |  |
| Min, Max | 19·2, 65·4 | 18·0, 70·8 |  | 20·6, 61·8 | 18·9, 63·7 |  | 18·4, 63·7 | 18·2, 69·7 |  |
| Age group, n(%) |  |  | 0·5519 |  |  | 0·3157 |  |  | 0·9830 |
| 18-59 years | 296 (98·34) | 292 (97·66) |  | 299 (99·67) | 297 (99·00) |  | 293 (98·32) | 297 (98·34) |  |
| ≥60 years | 5 (1·66) | 7 (2·34) |  | 1 (0·33) | 3 (1·00) |  | 5 (1·68) | 5 (1·66) |  |
| Sex, n(%) |  |  | 0·6146 |  |  | 0·5026 |  |  | 0·3923 |
| Male | 261 (86·71) | 255 (85·28) |  | 283 (94·33) | 279 (93·00) |  | 258 (86·58) | 254 (84·11) |  |
| Female | 40 (13·29) | 44 (14·72) |  | 17 (5·67) | 21 (7·00) |  | 40 (13·42) | 48 (15·89) |  |
| Height (cm) |  |  |  |  |  |  |  |  |  |
| Mean (SD) | 167·52 (7·87) | 167·61 (8·99) | 0·8938 | 169·93 (7·97) | 169·17 (7·79) | 0·2389 | 170·51 (8·25) | 169·47 (8·89) | 0·1400 |
| Median | 168·00 | 168·00 |  | 170·00 | 169·00 |  | 171·00 | 171·00 |  |
| Min, Max | 147·00, 190·00 | 125·00, 191·00 |  | 110·00, 187·00 | 143·00, 189·00 |  | 143·00, 190·50 | 141·00, 192·00 |  |
| Weight (kg) |  |  |  |  |  |  |  |  |  |
| Mean (SD) | 71·02 (14·22) | 71·96 (14·17) | 0·4165 | 76·10 (13·27) | 74·11 (13·22) | 0·0665 | 77·12 (15·56) | 76·46 (14·41) | 0·5894 |
| Median | 70·00 | 70·70 |  | 75·00 | 73·00 |  | 76·00 | 75·50 |  |
| Min, Max | 42·00, 133·00 | 44·10, 126·00 |  | 46·80, 119·00 | 43·00, 128·00 |  | 43·50, 132·70 | 43·00, 127·00 |  |

**Figure 1:**
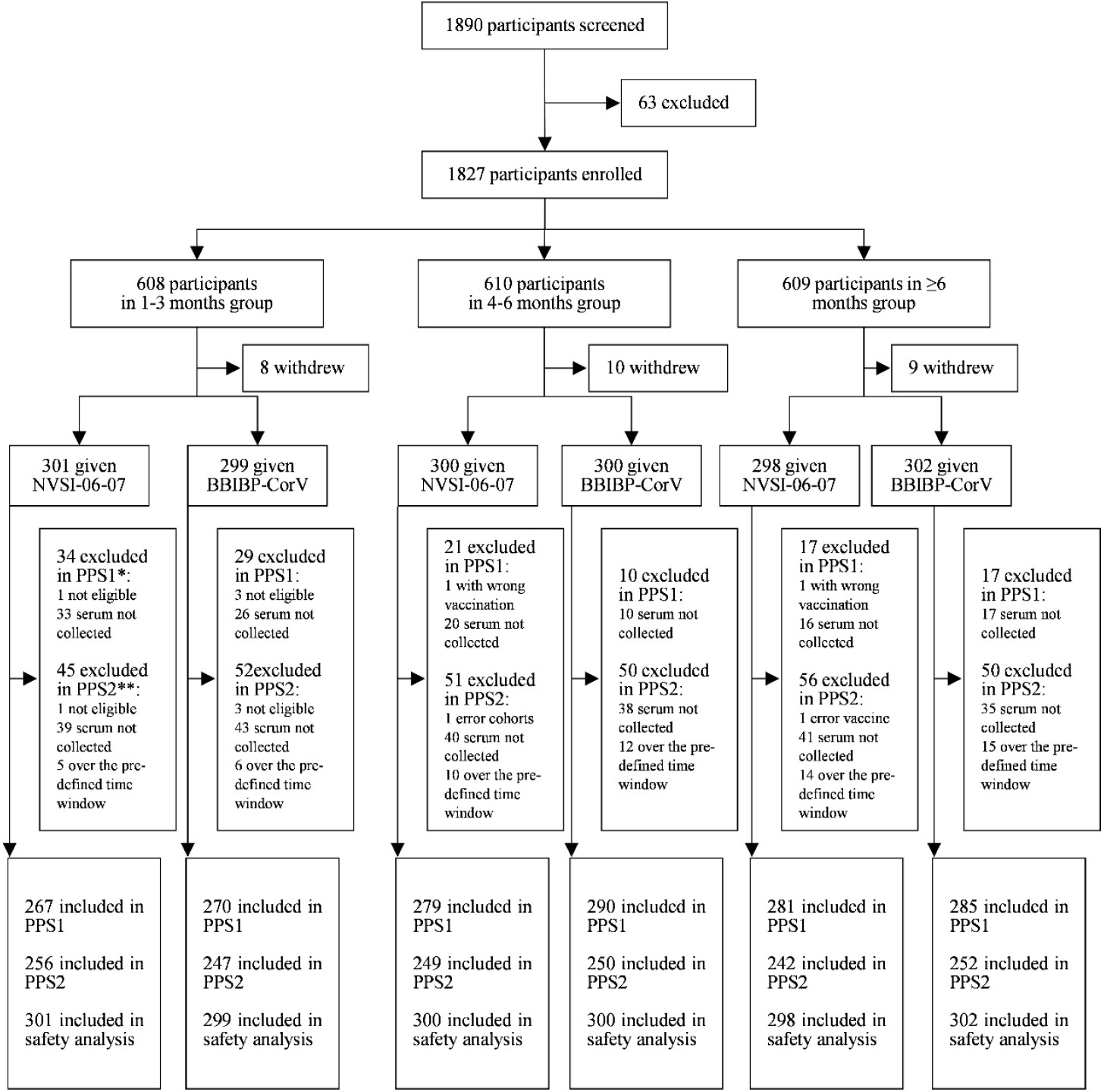
Trial profile. *PPS1: per-protocol analysis of immunogenicity on day 14 post booster vaccination; **PPS2: per-protocol analysis of immunogenicity on day 28 post booster vaccination; The sera from all the participants in PPS2 were used to evaluate the neutralizing antibody titers. 255 participants in the 1-3 months group receiving NVSI-06-07 boost, 241 in the ≥6 months group receiving NVSI-06-07, and 251 in the ≥6 months group receiving BBIBP-CorV were used to detect the RBD-binging IgG titers.

For safety analysis, four cases of SAEs were reported within 30 days post-boosting, two of which occurred in homologous booster group, and the other two was reported in heterologous booster group. None of these SAEs was related to the tested vaccines. Besides, no AESI was reported. The overall occurrence of adverse reactions was low in both the heterologous and homologous booster vaccinations. The most frequent adverse reactions were grades 1 (mild) or 2 (moderate) in severity (Figures 2 and 3, and Appendix Tables A2 and A3). Among participants boosted with NVSI-06-07, 184 (20·47%) reported at least one adverse reaction within 30 days post-boosting. And for the groups boosted with a third dose of BBIBP-CorV, the total number of participants reporting any adverse reaction was 177 (19·64%). No statistically significant difference was observed in the occurrence of adverse reactions between these two groups (*P*=0·6805) (Appendix Table A3).

**Figure 2.**
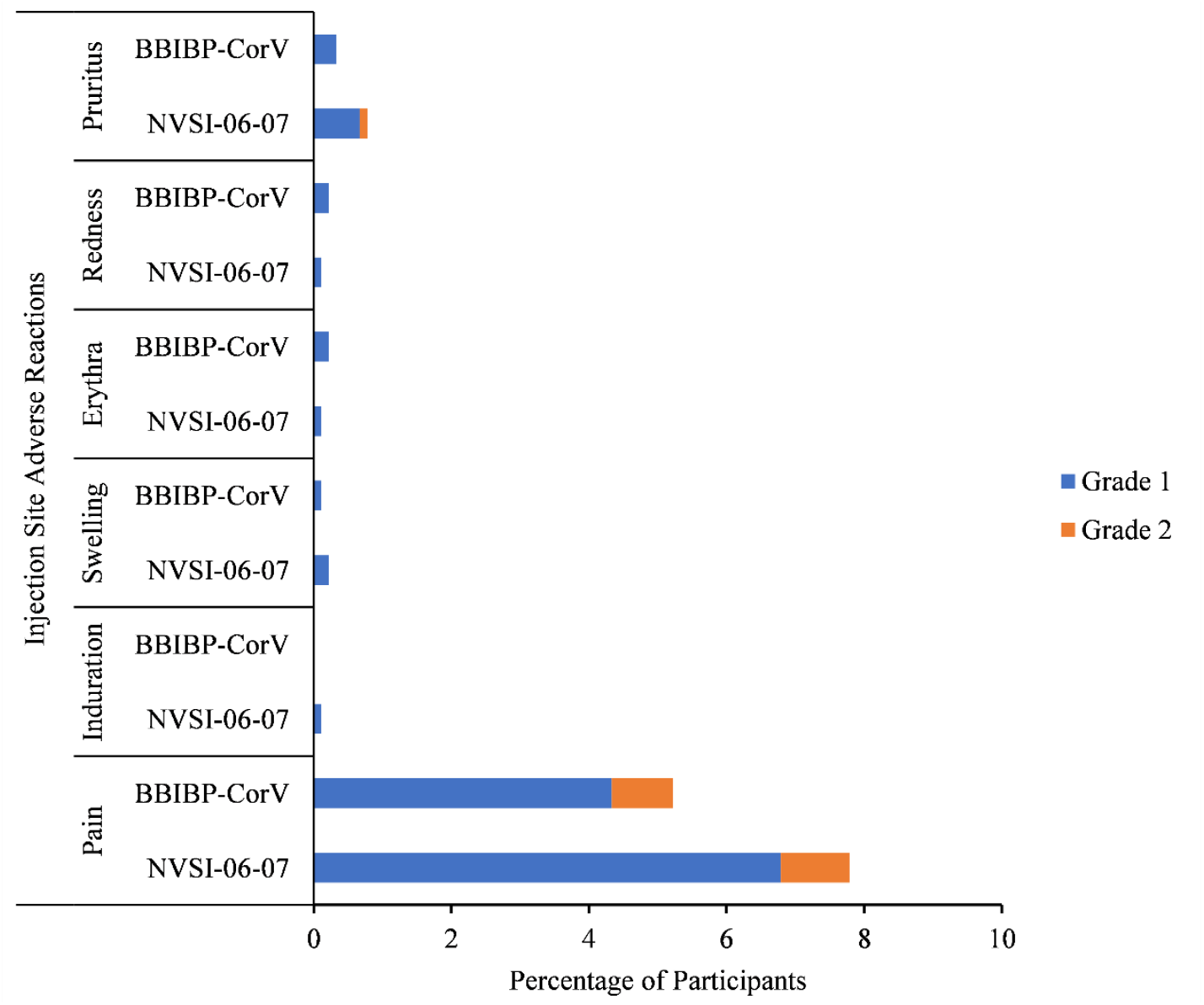
Injection site adverse reactions reported within 30 days after injection of NVSI-06-07 or BBIBP-CorV. Adverse reactions are graded according to the scale issued by the China National Medical Products Administration (NMPA). Grade 1 is mild and grade 2 is moderate.

**Figure 3.**
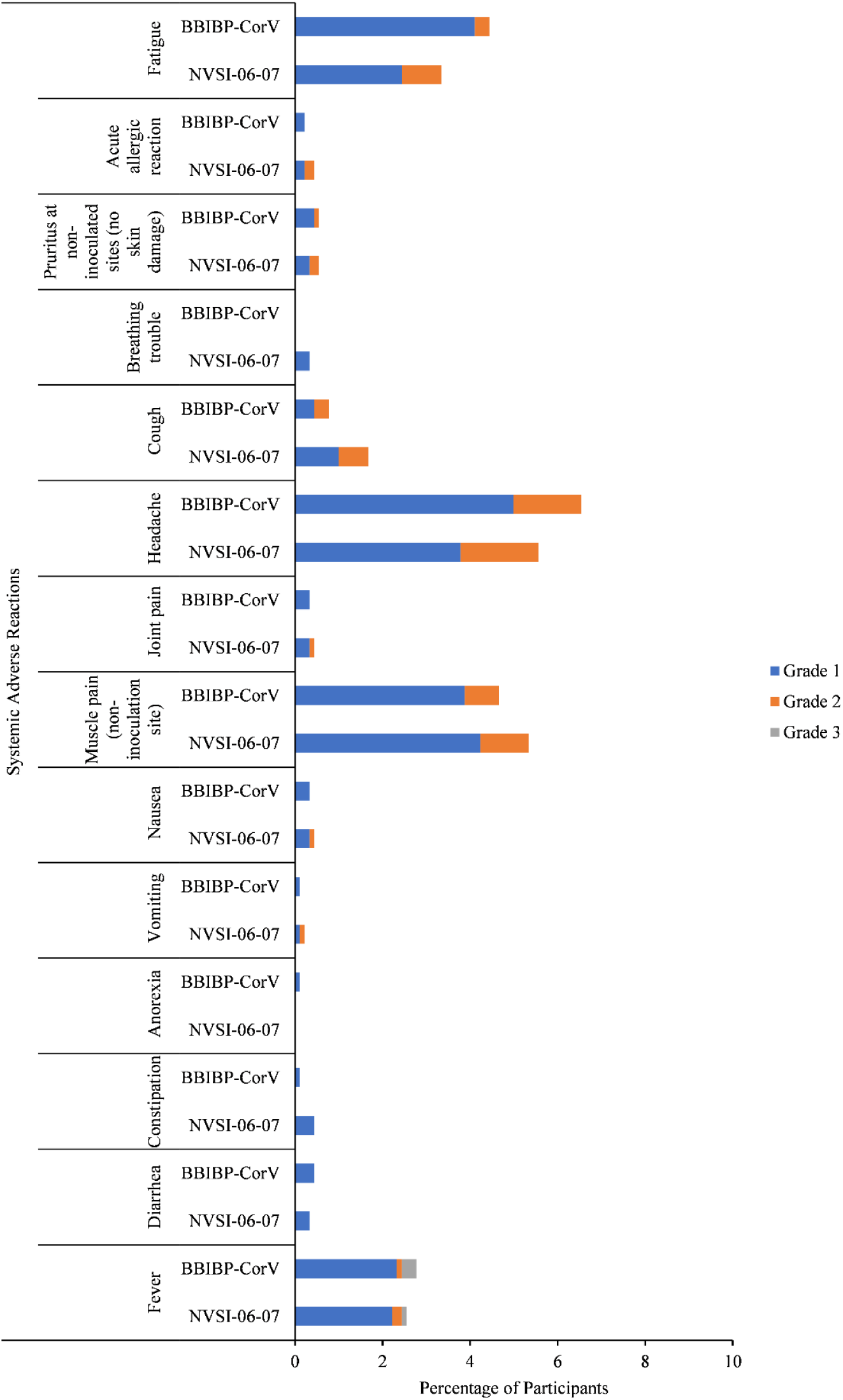
Systemic adverse reactions reported within 30 days after injection of NVSI-06-07 or BBIBP-CorV. Adverse reactions are graded according to the scale issued by the China National Medical Products Administration (NMPA). Grade 1 is mild, grade 2 is moderate, and grade 3 is severe.

The number of individuals reporting any unsolicited adverse event relevant to vaccination was 57 (6·34%) and 58 (6·44%) in heterologous and homologous boosting groups, respectively, within 30 days after booster vaccination (*P*=1·0000) (Appendix Table A3). These reported unsolicited adverse reactions were all ranked as grades 1 or 2. For solicited adverse reactions collected within 30 days post-boosting, most of the local and systemic adverse reactions were graded as 1 (mild) or 2 (moderate) in both heterologous and homologous boosting groups, except for grade 3 systemic fever reported by 1 participant (0·11%) in heterologous boosting group and 3participants (0·33%) in homologous boosting group (Figures 2 and Appendix Table A3). Most of the solicited adverse reactions occurred within 7 days (Appendix Table A2). The most common injection site adverse reaction within 30 days was pain, reported in 70 (7·79%) subjects in the NVSI-06-07 boosting recipients and 47 (5·22%) in the BBIBP-CorV boosting recipients. There was a statistically difference in the pain of grade 1 between these two groups (*P*=0·0237). The most common systemic adverse reactions were headaches, muscle pain (non-inoculation site), fatigues and fever, which were reported in 49 (5·45%), 48 (5·34%), 30 (3·34%) and 23 (2·56%) participants in NVSI-06-07 boosting recipients, and 59 (6·55%), 41 (4·55%), 40 (4·44%) and 24 (2·66%) in the BBIBP-CorV boosting recipients. No statistically significant differences were observed in systemic adverse reactions between the heterologous and homologous boosting groups (all *P*>0·05) (Figure 3 and Appendix Table A3).

For immunogenicity, the serologic RBD-specific IgG titers were detected before and after boost vaccination by using ELISA kits to assess the antibody responses. The baseline IgG levels in the enrolled participants were firstly detected, as shown in Table 2. There was no difference in the baseline IgG levels between participants assigned to heterologous and homologous boosting groups. On 14 days after boosting, notable increases were observed in IgG titers. In homologous BBIBP-CorV booster group, the seroconversion rates were 23·70% (95% CI,18·76%-29·24%), 25·17% (20·28%-30·58%) and 36·14% (30·56%-42·01%) for 1-3-month, 4-6-month and ≥6-month boosting-interval groups, respectively, whereas in heterologous NVSI-06-07 booster group, the seroconversion rates were 93·26% (95%CI, 89·55%-95·96%), 90·32% (86·23%-93·53%) and 85·77% (81·12%-89·63%), respectively (Table 2). Significantly higher seroconversion rates (*P*<0·0001) were elicited by heterologous boosting than by homologous vaccination (Table 2). For participants receiving the homologous boost, IgG GMTs increased from baseline by 2·76-fold (95%CI, 2·39-3·17) in 1-3-month boosting-interval group, 2·63-fold (95% CI, 2·26-3·06) in 4-6-month group and 4·71-fold (3·77-5·89) in ≥6-month group, respectively. Notedly, in participants receiving the heterologous boost with NVSI-06-07, IgG GMTs demonstrated a 43·41-fold (95%CI, 36·54-51·56), 44·68-fold (36·79-54·26) and 57·56-fold (44·72-74·07) of increases in the three groups with different boosting intervals, respectively (Table 2). IgG responses boosted by NVSI-06-07 were much higher (*P*<0.0001) than those by BBIBP-CorV (Table 2). Similar results were observed on day 28 post-boosting. Seroconversion rates were 84·23%-92·94% in different groups receiving heterologous boosting, which were significantly higher than those in participants receiving homologous boosting (17·60%-29·48%), as shown in Table 2. A similar increasing trend was observed in IgG GMTs, which were 1·97-3·57 folds in the groups receiving homologous prime-boost vaccination and 30·99-41·68 folds in the groups receiving heterologous prime-boost vaccination. On day 28 after boost, both seroconversion rates and IgG GMTs induced by the heterologous boost vaccination were significantly higher (*P*<0·0001) than those induced by the homologous boost vaccination (Table 2).

**Table 2:** RBD-specific IgG response results (PPS)

|  | 14 days after boosting |  |  | 28 days after boosting |  |  |
| --- | --- | --- | --- | --- | --- | --- |
|  | NVSI-06-07 | BBIBP-CorV | P value | NVSI-06-07 | BBIBP-CorV | P value |
| **1-3 months** |  |  |  |  |  |  |
| N(missing) | 267 (0) | 270 (0) |  | 255 (0) | 247 (0) |  |
| Pre-booster antibody GMT (95%CI) | 110·14 (93·50, 129·74) | 109·50 (94·24, 127·24) | 0·9591 | 106·64 (90·17, 126·10) | 106·30 (90·89, 124·33) | 0·9785 |
| Post-booster antibody GMT (95%CI) | 4780·76 (4106·48, 5565·76) | 301·71 (274·96, 331·07) |  | 3304·37 (2870·60, 3803·68) | 254·79 (231·81, 280·04) |  |
| Post-booster adjusted antibody GMT (95%CI) | 4776·34 (4255·49, 5360·94) | 301·99 (269·23, 338·73) |  | 3302·54 (2974·17, 3667·17) | 254·93 (229·20, 283·55) |  |
| Two groups ratio of adjusted GMT (95% CI) <sup>[1]</sup> | 15·82 (13·44, 18·61) |  | <0·0001<sup>[2]</sup> | 12·95 (11·16, 15·04) |  | <0·0001<sup>[2]</sup> |
| Rate of seroconversion* n (%) | 249 (93·26) | 64 (23·70) |  | 237 (92·94) | 54 (21·86) |  |
| 95%CI (%) | 89·55, 95·96 | 18·76, 29·24 |  | 89·07, 95·76 | 16·87, 27·54 |  |
| Two groups rate difference (%, 95%CI<sup>[3]</sup>) | 69·55 (63·66,75·45) |  | <0·0001 | 71·08 (65·04,77·12) |  | <0·0001 |
| Post-booster antibody GMT growth folds (95%CI) | 43·41 (36·54, 51·56) | 2·76(2·39, 3·17) | <0·0001 | 30·99 (26·47, 36·27) | 2·40 (2·09, 2·75) | <0·0001 |
| **4-6 months** |  |  |  |  |  |  |
| N(missing) | 279 (0) | 290 (0) |  | 249 (0) | 250 (0) |  |
| Pre-booster antibody GMT (95%CI) | 152·16 (127·35, 181·81) | 170·87 (142·36, 205·09) | 0·3714 | 150·77 (124·60, 182·44) | 185·48 (152·36, 225·80) | 0·1370 |
| Post-booster antibody GMT (95%CI) | 6798·51 (5985·06, 7722·52) | 448·93 (401·59, 501·84) |  | 4925·67 (4361·44, 5562·89) | 364·90 (324·38, 410·49) |  |
| Post-booster adjusted antibody GMT (95%CI) | 6901·42 (6174·88, 7713·43) | 442·48 (396·75, 493·49) |  | 5074·81 (4564·26, 5642·48) | 354·22 (318·65, 393·76) |  |
| Two groups ratio of adjusted GMT (95% CI) <sup>[1]</sup> | 15·60 (13·35, 18·23) |  | <0·0001<sup>[2]</sup> | 14·33 (12·33, 16·64) |  | <0·0001<sup>[2]</sup> |
| Rate of seroconversion* n (%) | 252 (90·32) | 73 (25·17) |  | 223 (89·56) | 44 (17·60) |  |
| 95%CI (%) | 86·23, 93·53 | 20·28, 30·58 |  | 85·08, 93·06 | 13·09, 22·90 |  |
| Two groups rate difference (%, 95%CI<sup>[3]</sup>) | 65·15 (59·07,71·23) |  | <0·0001 | 71·96 (65·90,78·02) |  | <0·0001 |
| Post-booster antibody GMT growth folds (95%CI) | 44·68 (36·79, 54·26) | 2·63 (2·26, 3·06) | <0·0001 | 32·67 (26·94, 39·62) | 1·97 (1·69, 2·29) | <0·0001 |
|  | NVSI-06-07 | BBIBP-CorV | P value | NVSI-06-07 | BBIBP-CorV | P value |
| **≥6 months** |  |  |  |  |  |  |
| N(missing) | 281 (0) | 285 (0) |  | 241 (0) | 251 (0) |  |
| Pre-booster antibody GMT (95%CI) | 104·94 (82·03, 134·24) | 125·99 (98·29, 161·50) | 0·3039 | 114·54 (88·18, 148·78) | 132·80 (102·75, 171·63) | 0·4268 |
| Post-booster antibody GMT (95%CI) | 6039·76 (5238·91, 6963·03) | 593·53 (519·22, 678·47) |  | 4774·32 (4157·08, 5483·21) | 473·53 (410·98, 545·60) |  |
| Post-booster adjusted antibody GMT (95%CI) | 6148·08 (5399·43, 7000·53) | 583·21 (512·67, 663·47) |  | 4846·54 (4249·18, 5527·88) | 466·76 (410·31, 530·97) |  |
| Two groups ratio of adjusted GMT (95% CI) <sup>[1]</sup> | 10·54 (8·78, 12·66) |  | <0·0001<sup>[2]</sup> | 10·38 (8·64, 12·48) |  | <0·0001<sup>[2]</sup> |
| Rate of seroconversion* n (%) | 241 (85·77) | 103 (36·14) |  | 203 (84·23) | 74 (29·48) |  |
| 95%CI (%) | 81·12, 89·63 | 30·56, 42·01 |  | 79·01, 88·59 | 23·91, 35·54 |  |
| Two groups rate difference (%, 95%CI<sup>[3]</sup>) | 49·62 (42·71, 56·54) |  | <0·0001 | 54·75 (47·47, 62·03) |  | <0·0001 |
| Post-booster antibody GMT growth folds (95%CI) | 57·56 (44·72, 74·07) | 4·71 (3·77, 5·89) | <0·0001 | 41·68 (32·05, 54·22) | 3·57 (2·84, 4·48) | <0·0001 |

Notes: [1] Two groups ratio of Post-booster adjusted GMT was “NVSI-06-07/ BBIBP-CorV”, and the non-inferiority threshold of ratio between groups 14 days/28 days in the protocol was set to 0·67.
[2] Covariance analysis adjusted model was used to calculate Least squares value and P value.
[3] Rate difference= (NVSI-06-07) - (BBIBP-CorV), Rate difference and 95%CI were estimated by CMH method considering stratification factors.
\* Seroconversion was defined as more than 4-fold rise from baseline in IgG titer.

The immunogenic superiority of heterologous NVSI-06-07 booster to homologous BBIBP-CorV booster was further confirmed by neutralizing antibody response measured with live-virus neutralization assays. Before booster vaccination, most of the participants had detectable neutralizing activities against prototype SARS-CoV-2 and showed a comparable level between two boosting groups in the pre-booster neutralizing antibodies. The pre-booster neutralizing antibody GMT of participants in the group of over-6-month boosting-interval was about half of the values in the 3-6-month group, indicating wanning of neutralizing antibody responses over time (Table 3). On day 14 post-boosting, the neutralizing antibody titers against prototype SARS-CoV-2 live virus were significantly improved in both the heterologous and homologous boosting recipients. In homologous boosting participants, the seroconversion rates in 1-3-month, 4-6-month and ≥6-month boosting-interval groups were 39·26% (95%CI, 33·40%-45·36%), 26·90% (21·88%-32·39%) and 52·98% (47·01%-58·89%), respectively, whereas those were 81·65% (76·47%-86·10%), 86·38% (81·79%-90·18%) and 86·83% (82·31%-90·56%) for the heterologous boost (Table 3). The seroconversion rates induced by heterologous boost were significantly higher (*P*<0·0001) than those induced by homologous boost (Table 3). Compared with the pre-boosting baseline level, the homologous boost vaccination elicited 3·41-fold (95%CI, 2·90-4·00) higher neutralizing GMTs against prototype SARS-CoV-2 in 1-3-month boosting-interval group, 2·58-fold (95% CI, 2·21-3·00) higher in 4-6-month group and 7·36-fold (95%CI, 6·11-8·86) higher in ≥6-month group, respectively. A more remarkable improvement of neutralizing antibody responses was observed by heterologous boost vaccination against prototype live virus, in which average neutralizing GMTs increased by 13·95-fold (95%CI, 12·01-16·20), 16·45-fold (14·10-19·19) and 35·86-fold (29·44-43·67) for the three groups (Table 3). On day 28 post-boosting, live-virus neutralizing antibody responses were further improved in both homologous and heterologous boosting groups. By homologous boosting, seroconversion rates were further increased to 59·92% (95%CI, 53·52%-66·08%), 36·80%(30·81%-43·11%) and 81·75% (76·41%-86·31%) in the 1-3-month, 4-6-month and ≥6-month boosting-interval groups, respectively. Much higher seroconversion rates were obtained by heterologous boost, which reached at 90·63% (95%CI, 86·37%-93·90%), 89·96% (85·54%-93·40%) and 97·52% (94·68%-99·08%) in the three groups, respectively. On day 28 post-boost, the increases from baseline in neutralizing GMTs of heterologous prime-boost vaccination were also significantly higher than those of homologous vaccination. By homologous boosting, neutralizing GMTs improved by 7·08-fold (95%CI, 5·91-8·48), 4·20-fold (3·57-4·94) and 16·78-fold (13·51-20·83) in the three groups, respectively, whereas 21·01-fold (95%CI, 18·01-24·52), 23·10-fold (19·44-27·44) and 63·85-fold (52·15-78·18) of increase were obtained by heterologous boost (Table 3). Both on day 14 and day 28 post-boosting, neutralizing antibody levels improved by heterologous booster were much higher (*P*<0·0001) than those by homologous booster, indicating that NVSI-06-07 is immunologically preferred as a booster choice over BBIBP-CorV (Table 3). Comparison among three groups with different prime-boosting intervals showed that the ≥6 months groups have the largest increase in neutralizing GMTs both for heterologous and homologous boosts, and thus the booster vaccination with over 6 months intervals was optimal.

**Table 3:** Live-virus neutralizing antibody response results (PPS)

|  | 14 days after boosting |  |  | 28 days after boosting |  |  |
| --- | --- | --- | --- | --- | --- | --- |
|  | NVSI-06-07 | BBIBP-CorV | P value | NVSI-06-07 | BBIBP-CorV | P value |
| **1-3 months** |  |  |  |  |  |  |
| N(missing) | 267 (0) | 270 (0) |  | 256 (0) | 247 (0) |  |
| Pre-booster antibody GMT (95%CI) | 95·71 (81·88, 111·88) | 86·93 (73·84, 102·33) | 0·4018 | 93·44 (79·36, 110·02) | 83·63 (70·42, 99·31) | 0·3571 |
| Post-booster antibody GMT (95%CI) | 1335·43 (1152·56, 1547·31) | 296·20 (266·22, 329·55) |  | 1963·31 (1713·49, 2249·55) | 592·12 (528·52, 663·38) |  |
| Post-booster adjusted antibody GMT (95%CI) | 1313·12 (1169·39, 1474·52) | 301·17 (268·38, 337·97) |  | 1933·53 (1722·58, 2170·32) | 601·57 (534·82, 676·66) |  |
| Two groups ratio of adjusted GMT (95% CI) <sup>[1]</sup> | 4·36 (3·70, 5·13) |  | <0·0001<sup>[2]</sup> | 3·21(2·73, 3·79) |  | <0·0001<sup>[2]</sup> |
| Rate of seroconversion* n (%) | 218 (81·65) | 106 (39·26) |  | 232 (90·63) | 148 (59·92) |  |
| 95%CI (%) | 76·47, 86·10 | 33·40, 45·36 |  | 86·37, 93·90 | 53·52, 66·08 |  |
| Two groups rate difference (%, 95%CI<sup>[3]</sup>) | 42·39 (34·94, 49·84) |  | <0·0001 | 30·71 (23·63, 37·78) |  | <0·0001 |
| Post-booster antibody GMT growth folds (95%CI) | 13·95 (12·01, 16·20) | 3·41 (2·90, 4·00) | <0·0001 | 21·01 (18·01, 24·52) | 7·08 (5·91, 8·48) | <0·0001 |
| **4-6 months** |  |  |  |  |  |  |
| N(missing) | 279 (0) | 290 (0) |  | 249 (0) | 250 (0) |  |
| Pre-booster antibody GMT (95%CI) | 110·22 (93·72, 129·63) | 127·18 (108·73, 148·76) | 0·2121 | 109·41 (91·81, 130·39) | 138·25 (117·12, 163·19) | 0·0569 |
| Post-booster antibody GMT (95%CI) | 1812·82 (1604·36, 2048·37) | 327·81 (297·72, 360·93) |  | 2527·18 (2213·43, 2885·41) | 580·70 (518·66, 650·16) |  |
| Post-booster adjusted antibody GMT (95%CI) | 1848·96 (1670·38, 2046·62) | 321·64 (291·14, 355·33) |  | 2612·33 (2332·20, 2926·09) | 561·85 (501·72, 629·19) |  |
| Two groups ratio of adjusted GMT (95% CI) <sup>[1]</sup> | 5·75 (4·99, 6·63) |  | <0·0001<sup>[2]</sup> | 4·65 (3·96, 5·46) |  | <0·0001<sup>[2]</sup> |
| Rate of seroconversion* n (%) | 241 (86·38) | 78 (26·90) |  | 224 (89·96) | 92 (36·80) |  |
| 95%CI (%) | 81·79, 90·18 | 21·88, 32·39 |  | 85·54, 93·40 | 30·81, 43·11 |  |
| Two groups rate difference (%, 95%CI<sup>[3]</sup>) | 59·48 (52·98,65·98) |  | <0·0001 | 53·16 (46·11,60·21) |  | <0·0001 |
| Post-booster antibody GMT growth folds (95%CI) | 16·45 (14·10, 19·19) | 2·58 (2·21, 3·00) | <0·0001 | 23·10 (19·44, 27·44) | 4·20 (3·57, 4·94) | <0·0001 |
|  | NVSI-06-07 | BBIBP-CorV | P value | NVSI-06-07 | BBIBP-CorV | P value |
| **≥6 months** |  |  |  |  |  |  |
| N(missing) | 281 (0) | 285 (0) |  | 242 (0) | 252 (0) |  |
| Pre-booster antibody GMT (95%CI) | 53·17 (43·28, 65·32) | 61·07 (50·26, 74·20) | 0·3363 | 59·05 (47·80, 72·95) | 64·92 (53·10, 79·38) | 0·5224 |
| Post-booster antibody GMT (95%CI) | 1906·60 (1651·75, 2200·77) | 449·30 (406·41, 496·72) |  | 3770·62 (3263·18, 4356·98) | 1089·23 (959·93, 1235·95) |  |
| Post-booster adjusted antibody GMT (95%CI) | 1937·97 (1728·18, 2173·23) | 442·13 (394·58, 495·40) |  | 3806·74 (3341·56, 4336·68) | 1079·31(949·89, 1226·35) |  |
| Two groups ratio of adjusted GMT (95% CI) <sup>[1]</sup> | 4·38 (3·73, 5·15) |  | <0·0001<sup>[2]</sup> | 3·53 (2·94, 4·23) |  | <0·0001<sup>[2]</sup> |
| Rate of seroconversion* n (%) | 244 (86·83) | 151 (52·98) |  | 236 (97·52) | 206 (81·75) |  |
| 95%CI (%) | 82·31, 90·56 | 47·01, 58·89 |  | 94·68, 99·08 | 76·41, 86·31 |  |
| Two groups rate difference (%, 95%CI<sup>[3]</sup>) | 33·85 (26·84,40·87) |  | <0·0001 | 15·77 (10·62,20·93) |  | <0·0001 |
| Post-booster antibody GMT growth folds (95%CI) | 35·86 (29·44, 43·67) | 7·36 (6·11, 8·86) | <0·0001 | 63·85 (52·15, 78·18) | 16·78 (13·51, 20·83) | <0·0001 |

Notes: [1] Two groups ratio of Post-booster adjusted GMT was “NVSI-06-07/ BBIBP-CorV”, and the non-inferiority threshold of ratio between groups 14 days/28 days in the protocol was set to 0·67.
[2] Covariance analysis adjusted model was used to calculate Least squares value and P value.
[3] Rate difference= (NVSI-06-07) - (BBIBP-CorV), Rate difference and 95%CI were estimated by CMH method considering stratification factors.
\* Seroconversion was defined as more than 4-fold rise form baseline in neutralizing antibody titer.

Owing to much higher transmissibility than other VOCs, the Omicron variant has rapidly spread around the world. Worryingly, Omicron has been demonstrated with substantially improved immune-escape capability by many preliminary studies,^14-16^ which raised serious concerns on the effectiveness of available COVID-19 vaccines. In this study, serum samples of 192 participants with consecutive enrollment numbers in ≥6-month boosting-interval group (half boosted with homologous vaccination and the other half boosted with heterologous vaccination) were used to evaluate the neutralizing sensitivities to the Omicron variant using live-virus neutralization assays. In participants boosted with a third dose of BBIBP-CorV, neutralizing antibody GMT against Omicron was substantially reduced by 11·32 folds compared with that against prototype SARS-CoV-2 strain, implying substantial escape of the Omicron variant from the antibody neutralization response elicited by BBIBP-CorV. By comparison, in participants receiving heterologous boost of NVSI-06-07, neutralizing antibody GMT against Omicron only declined by 6·62 folds, as shown in Table 4. Neutralizing antibody GMT against Omicron elicited by heterologous boost was 292·53 (95%CI, 222·81-384·07), which was significantly higher than 37·91 (95% CI, 30·35-47·35) induced by homologous boost. Heterologous prime-booster vaccination with BBIBP-CorV followed by NVSI-06-07 demonstrated much more robust neutralizing activities against Omicron compared with homologous prime-boost vaccination with three doses of BBIBP-CorV.

**Table 4:**
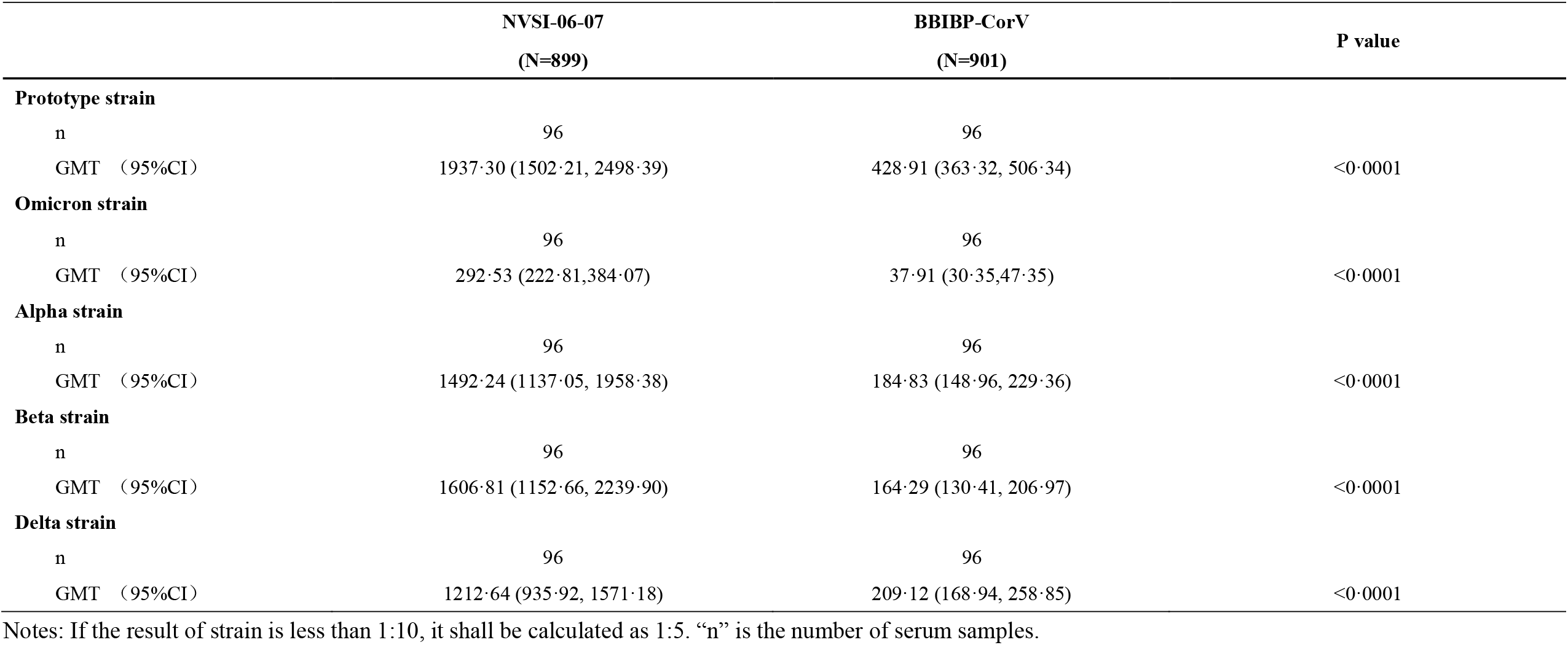
Live-virus neutralizing antibody responses against main SARS-CoV-2 VOCs.

|  | **NVSI-06-07**<br>(N=899) | **BBIBP-CorV**<br>(N=901) | **P value** |
| --- | --- | --- | --- |
| **Prototype strain** |  |  |  |
| n | 96 | 96 |  |
| GMT (95%CI) | 1937.30 (1502.21, 2498.39) | 428.91 (363.32, 506.34) | <0.0001 |
| **Omicron strain** |  |  |  |
| n | 96 | 96 |  |
| GMT (95%CI) | 292.53 (222.81,384.07) | 37.91 (30.35,47.35) | <0.0001 |
| **Alpha strain** |  |  |  |
| n | 96 | 96 |  |
| GMT (95%CI) | 1492.24 (1137.05, 1958.38) | 184.83 (148.96, 229.36) | <0.0001 |
| **Beta strain** |  |  |  |
| n | 96 | 96 |  |
| GMT (95%CI) | 1606.81 (1152.66, 2239.90) | 164.29 (130.41, 206.97) | <0.0001 |
| **Delta strain** |  |  |  |
| n | 96 | 96 |  |
| GMT (95%CI) | 1212.64 (935.92, 1571.18) | 209.12 (168.94, 258.85) | <0.0001 |

Notes: If the result of strain is less than 1:10, it shall be calculated as 1:5. “n” is the number of serum samples.

We also evaluated the immune efficacy of booster vaccinations against other SARS-CoV-2 VOCs, including Alpha, Beta and Delta. After boosting with BBIBP-CorV, the neutralizing antibody GMTs against Alpha, Beta and Delta showed 2·32, 2·61 and 2·05 folds decrease compared to that against prototype strain. All these three VOCs exhibited less sensitivities to sera neutralization, among which Beta variant showed the largest reduction in neutralization sensitivity. Our studies were consistent with previously reported results.^17-19^ By comparison, the sera from the participants boosted with NVSI-06-07 showed only 1·30, 1·21 and 1·60 folds reduction in neutralization of the Alpha, Beta and Delta variants, respectively. By heterologous booster vaccination, neutralizing antibody GMTs against these three VOCs were 1492·24 (95% CI,1137·05-1958·38), 1606·81 (1152·66-2239·90) and 1212·64 (935·92-1571·18), respectively, whereas the GMTs by homologous boost were 184·83 (148·96-229·36), 164·29 (130·41-206·97) and 209·12 (168·94-258·85), respectively (Table 4). The heterologous boost elicited much higher neutralizing activities against the tested VOCs.

## DISCUSSION

Findings from this trial showed that both heterologous boost with NVSI-06-07 and homologous boost with BBIBP-CorV were immunogenic in the BBIBP-CorV recipients, but the immune efficacy of heterologous boost was much greater than that of homologous boost. The fold increases in both IgG and neutralizing antibody GMTs from the corresponding baseline were significantly higher after heterologous boost than those after homologous boost. Especially, for adults primed with BBIBP-CorV over 6 months ago, a 63·85-fold increase in neutralizing antibody GMTs was obtained by heterologous boost, in comparison to 16·78 folds by homologous boost. Compared with the peak value of neutralizing antibody titers primed with two doses of BBIBP-CorV as reported in the previous literature^20^, neutralizing GMTs boosted by NVSI-06-07 were improved 6·94-13·34 folds on 28 days post-boosting. In comparison, homologous vaccination with a third dose of BBIBP-CorV only induced 2·09-3·85 folds increase over the peak value. Based on the comparison of the immune enhancements among groups with different prime-boosting intervals, our study suggested that the prime-booster vaccination with an over 6 months interval was optimal. The overall occurrence of adverse reactions was low in both heterologous and homologous boost vaccinations. Most of reported adverse reactions were graded as mild or moderate with the most common symptoms of injection-site pain, headaches, muscle pain (non-inoculation site), fatigues and fever. Reactogenicity of the booster vaccinations was similar to that of the priming vaccinations described in the previously published literatures,^20^ and there was no obvious difference in overall safety between heterologous and homologous boosts. The heterologous prime-boost combinations among viral vector COVID-19 vaccines, inactivated vaccines and mRNA vaccines have been proved to be able to significantly improve immune responses, and heterologous boost was more immunogenic than homologous boost.^8-10^ To our knowledge, this is the first reported result of a large-scale randomised, controlled clinical trial of heterologous boost with the recombinant subunit vaccine in the inactivated vaccine recipients. Our findings support the heterologous BBIBP-CorV/NVSI-06-07 prime-boost vaccination serving as one of the effective prime-boosting strategies to better combat SARS-CoV-2.

Our study showed that heterologous boost with NVSI-06-07 in persons primed by BBIBP-CorV not only substantially increased neutralization, but also improved the breadth of neutralizing response. Compared to homologous boost with a third dose of BBIBP-CorV, significantly higher neutralizing antibody responses against SARS-CoV-2 VOCs, including Omicron, Alpha, Beta and Delta, were achieved by heterologous boost with NVSI-06-07. The results were consistent with other studies.^11,12,21^ Currently, Omicron variant spreads rapidly around the world. However, Omicron-specific vaccine is still not available and other strategies are urgently needed to control the pandemic of this variant. Considering that BBIBP-CorV has been applied in large-scale populations and the BBIBP-CorV/NVSI-06-07 prime-booster vaccination can elicit a certain level of neutralizing antibodies against Omicron, this heterologous prime-booster vaccination might serve as a possible strategy combating Omicron.

Many studies have revealed that the levels of neutralizing antibody response were highly correlated with the real-world protection efficacy of the COVID-19 vaccines.^22-27^ According to the previously determined threshold indicative of reduced risks of symptomatic infection (506·00 BAU/mL),^27^ heterologous prime-boost vaccination of BBIBP-CorV combined with NVSI-06-07 might provide protective effects against SARS-CoV-2 in the real-world. But further studies are warranted to assess the long-term protection of this prime-booster strategy.

This study has limitations. First, for the volunteers enrolled in the trial, the number of men was much larger than that of women, and thus the data did not well represent the immune effects on women. Second, the proportion of older individuals aged ≥60 years in the participants was small, and the immune response was analyzed without taking into account different age ranges of the participants. Third, data on immune persistence of the booster vaccination is not yet available, and the long-term immunogenicity needs to be further studied.

In summary, heterologous booster vaccination with NVSI-06-07 in BBIBP-CorV recipients was well tolerated and immunogenic against not only SARS-CoV-2 prototype strain but also the VOCs including Omicron, which supported the approval of emergency use of this heterologous booster strategy.

## Data Availability

All data produced in the present work are contained in the manuscript

## Contributors

NA was the chief investigator. NA, QML, YTZ, XMY, YKY, JingZ^#^ and YK designed the trial and study protocol. QML, JingZ^#^, YL and JGS designed the recombinant vaccine NVSI-06-07. QML, YTZ and XMY were responsible for the organization and supervision of the project. QML, JingZ^#^, YL, XJG and XYM provided NVSI-06-07 vaccine. HW and JinZ provided the inactivated vaccine BBIBP-CorV. XMY, YTZ, IE and YKY contributed to project management. TY, YK, ML, LQ, WZ, PX, XZ, CQ, DYL and SSY participated in the implementation of the trial. HMM, ZNW and JLY conducted sample collection and processing. GW, KX, WL, JingZ^‡^, MY and SM carried out the serum tests. GW, KX, YL and FJS contributed to the development of serum testing method and manuscript preparation. SH, MSE and NL performed the clinical data gathering, analysis and operation. ZJ performed statistical analysis of the data. QML, JingZ^#^, YL, GW, YKY and KX performed data analysis and interpretation. JGS, JingZ^#^, YL, KX, SS and SSY contributed to the writing of the manuscript. QML and GW oversaw final manuscript preparation. All authors reviewed and approved the final manuscript.

## Declaration of interests

JingZ^#^, YL, YK, JGS, SS, SSY, NL, FJS, QML are employees of the National Vaccine and Serum Institute (NVSI). YKY, TY, ML, XZ, CQ, DYL, ZNW, JLY, LQ, YTZ, XMY are employees of the China National Biotec Group Company Limited. XJG, XYM, are employees of Lanzhou Institute of Biological Products Company Limited. HW, JinZ are employees of Beijing Institute of Biological Products Company Limited. ZJ is an employee of Beijing KeyTech Statistical Consulting Co.,Ltd. HMM is an employee of Union 71, United Arab Emirates. WZ, SM, IE, PX are employees of G42 Healthcare, IROS (Insights Research Organization & Solutions), United Arab Emirates. JingZ^#^, YL, JGS, SS, NL, QML are listed as inventors of the patent applications for the trimeric RBD-based vaccines (Application numbers: 202110348881.6, 202110464788.1 and 202110676901.2) and a patent (Application number: 202110676901.2) has been authorized. The other authors declare no competing interests.

## Data sharing

The clinical trial is still ongoing, and the individual participant data will be available when the trial is complete, upon requests to the corresponding author (QML). After proposals are approved, data can be shared through secure online platforms.

## Acknowledgements

We would like to thank Prof. Guoyong Yuan from the University of Hong Kong for providing SARS-CoV-2 Omicron virus applied in live-virus neutralization assay. We also thank Yao Tan and Xiang Feng Cong for their valuable help in the manuscript preparation.

**Appendix Table A1:** Baseline characteristic for the nationality of the participants (FAS)

|  | 1-3 months |  |  | 4-6 months |  |  | ≥6 months |  |  |
| --- | --- | --- | --- | --- | --- | --- | --- | --- | --- |
|  | NVSI-06-07<br>(N=301) | BBIBP-CorV<br>(N=299) | P value | NVSI-06-07<br>(N=300) | BBIBP-CorV<br>(N=300) | P value | NVSI-06-07<br>(N=298) | BBIBP-CorV<br>(N=302) | P value |
| **Countries, n(%)** |  |  |  |  |  |  |  |  |  |
| Afghanistan | 9 (2.99) | 15 (5.02) | 0.3109 | 2 (0.67) | 1 (0.33) | 0.2941 | 0 (0.00) | 1 (0.33) | 0.7785 |
| Bangladesh | 96 (31.89) | 76 (25.42) |  | 53 (17.67) | 49 (16.33) |  | 15 (5.03) | 15 (4.97) |  |
| Cameroon | 1 (0.33) | 2 (0.67) |  | 0 (0.00) | 0 (0.00) |  | 1 (0.34) | 1 (0.33) |  |
| Canada | 0 (0.00) | 1 (0.33) |  | 0 (0.00) | 0 (0.00) |  | 0 (0.00) | 0 (0.00) |  |
| Chad | 0 (0.00) | 0 (0.00) |  | 0 (0.00) | 0 (0.00) |  | 1 (0.34) | 0 (0.00) |  |
| Comorin | 1 (0.33) | 0 (0.00) |  | 0 (0.00) | 0 (0.00) |  | 0 (0.00) | 0 (0.00) |  |
| Egypt | 5 (1.66) | 8 (2.68) |  | 4 (1.33) | 8 (2.67) |  | 7 (2.35) | 13 (4.30) |  |
| United Arab Emirates | 0 (0.00) | 0 (0.00) |  | 2 (0.67) | 0 (0.00) |  | 15 (5.03) | 18 (5.96) |  |
| Ethiopia | 0 (0.00) | 0 (0.00) |  | 0 (0.00) | 0 (0.00) |  | 1 (0.34) | 0 (0.00) |  |
| Falkland Islands | 0 (0.00) | 0 (0.00) |  | 1 (0.33) | 0 (0.00) |  | 0 (0.00) | 0 (0.00) |  |
| Philippines | 30 (9.97) | 36 (12.04) |  | 12 (4.00) | 22 (7.33) |  | 26 (8.72) | 28 (9.27) |  |
| Ghana | 0 (0.00) | 0 (0.00) |  | 0 (0.00) | 1 (0.33) |  | 6 (2.01) | 5 (1.66) |  |
| India | 44 (14.62) | 46 (15.38) |  | 91 (30.33) | 84 (28.00) |  | 52 (17.45) | 47 (15.56) |  |
| Indonesia | 0 (0.00) | 0 (0.00) |  | 1 (0.33) | 0 (0.00) |  | 0 (0.00) | 0 (0.00) |  |
| Iran | 0 (0.00) | 1 (0.33) |  | 0 (0.00) | 0 (0.00) |  | 0 (0.00) | 0 (0.00) |  |
| Iraq | 1 (0.33) | 1 (0.33) |  | 0 (0.00) | 0 (0.00) |  | 0 (0.00) | 0 (0.00) |  |
| Jordan | 5 (1.66) | 11 (3.68) |  | 0 (0.00) | 0 (0.00) |  | 1 (0.34) | 0 (0.00) |  |
| Korea | 0 (0.00) | 0 (0.00) |  | 0 (0.00) | 0 (0.00) |  | 0 (0.00) | 0 (0.00) |  |
| Lebanon | 0 (0.00) | 0 (0.00) |  | 0 (0.00) | 0 (0.00) |  | 1 (0.34) | 0 (0.00) |  |
| Mauritania | 0 (0.00) | 0 (0.00) |  | 0 (0.00) | 0 (0.00) |  | 0 (0.00) | 2 (0.66) |  |
| Morocco | 5 (1.66) | 1 (0.33) |  | 4 (1.33) | 2 (0.67) |  | 8 (2.68) | 5 (1.66) |  |
|  | NVSI-06-07<br>(N=301) | BBIBP-CorV<br>(N=299) | P value | NVSI-06-07<br>(N=300) | BBIBP-CorV<br>(N=300) | P value | NVSI-06-07<br>(N=298) | BBIBP-CorV<br>(N=302) | P value |
| Nepal | 10 (3·32) | 16 (5·35) |  | 12 (4·00) | 21 (7·00) |  | 35 (11·74) | 36 (11·92) |  |
| Nigeria | 1 (0·33) | 0 (0·00) |  | 0 (0·00) | 1 (0·33) |  | 0 (0·00) | 0 (0·00) |  |
| Oman | 0 (0·00) | 3 (1·00) |  | 0 (0·00) | 0 (0·00) |  | 0 (0·00) | 0 (0·00) |  |
| Pakistan | 80 (26·58) | 66 (22·07) |  | 115 (38·33) | 105 (35·00) |  | 106 (35·57) | 104 (34·44) |  |
| Palestine | 4 (1·33) | 6 (2·01) |  | 0 (0·00) | 2 (0·67) |  | 0 (0·00) | 0 (0·00) |  |
| Sierra Leone | 0 (0·00) | 0 (0·00) |  | 0 (0·00) | 0 (0·00) |  | 1 (0·34) | 1 (0·33) |  |
| Sri Lanka | 0 (0·00) | 1 (0·33) |  | 1 (0·33) | 0 (0·00) |  | 0 (0·00) | 2 (0·66) |  |
| Sudan | 2 (0·66) | 3 (1·00) |  | 0 (0·00) | 1 (0·33) |  | 9 (3·02) | 8 (2·65) |  |
| Syria | 1 (0·33) | 0 (0·00) |  | 0 (0·00) | 0 (0·00) |  | 2 (0·67) | 0 (0·00) |  |
| Tanzania | 1 (0·33) | 0 (0·00) |  | 0 (0·00) | 0 (0·00) |  | 0 (0·00) | 0 (0·00) |  |
| Tunisia | 0 (0·00) | 0 (0·00) |  | 0 (0·00) | 0 (0·00) |  | 0 (0·00) | 2 (0·66) |  |
| Uganda | 5 (1·66) | 4 (1·34) |  | 2 (0·67) | 2 (0·67) |  | 8 (2·68) | 11 (3·64) |  |
| Yemen | 0 (0·00) | 0 (0·00) |  | 0 (0·00) | 1 (0·33) |  | 3 (1·01) | 3 (0·99) |  |
| Germany | 0 (0·00) | 1 (0·33) |  | 0 (0·00) | 0 (0·00) |  | 0 (0·00) | 0 (0·00) |  |
| Iberia | 0 (0·00) | 1 (0·33) |  | 0 (0·00) | 0 (0·00) |  | 0 (0·00) | 0 (0·00) |  |
| Other | 0 (0·00) | 0 (0·00) |  | 0 (0·00) | 0 (0·00) |  | 0 (0·00) | 0 (0·00) |  |

**Appendix Table A2:** Adverse reactions within 7 days of booster vaccination.

|  | NVSI-06-07<br>(N=899) | BBIBP-CorV<br>(N=901) | Total<br>(N=1800) | P value* |
| --- | --- | --- | --- | --- |
| **Solicited adverse reactions within 0-7 days** | 166 (18.46) | 151 (16.76) | 317 (17.61) | 0.3537 |
| **Injection site adverse reactions** | 73 (8.12) | 49 (5.44) | 122 (6.78) | 0.0245 |
| Pain | 70 (7.79) | 47 (5.22) | 117 (6.50) | 0.0280 |
| Induration | 1 (0.11) | 0 | 1 (0.06) | 0.4994 |
| Swelling | 2 (0.22) | 1 (0.11) | 3 (0.17) | 0.6244 |
| Erythra | 1 (0.11) | 2 (0.22) | 3 (0.17) | 1.0000 |
| Redness | 1 (0.11) | 2 (0.22) | 3 (0.17) | 1.0000 |
| Pruritus | 6 (0.67) | 2 (0.22) | 8 (0.44) | 0.1784 |
| **Systemic adverse reactions** | 120 (13.35) | 122 (13.54) | 242 (13.44) | 0.9449 |
| Fever | 18 (2.00) | 21 (2.33) | 39 (2.17) | 0.7466 |
| Diarrhea | 2 (0.22) | 4 (0.44) | 6 (0.33) | 0.6870 |
| Constipation | 4 (0.44) | 1 (0.11) | 5 (0.28) | 0.2177 |
| Anorexia | 0 | 1 (0.11) | 1 (0.06) | 1.0000 |
| Vomiting | 0 | 1 (0.11) | 1 (0.06) | 1.0000 |
| Nausea | 3 (0.33) | 3 (0.33) | 6 (0.33) | 1.0000 |
| Muscle pain (non-inoculation site) | 45 (5.01) | 41 (4.55) | 86 (4.78) | 0.6603 |
| Joint pain | 3 (0.33) | 3 (0.33) | 6 (0.33) | 1.0000 |
| Headache | 48 (5.34) | 56 (6.22) | 104 (5.78) | 0.4797 |
| Cough | 10 (1.11) | 5 (0.55) | 15 (0.83) | 0.2078 |
| Breathing trouble | 3 (0.33) | 0 | 3 (0.17) | 0.1244 |
| Pruritus at non-inoculated sites (no skin damage) | 4 (0.44) | 5 (0.55) | 9 (0.50) | 1.0000 |
| Acute allergic reaction | 3 (0.33) | 1 (0.11) | 4 (0.22) | 0.3740 |
| Fatigue | 27 (3.00) | 38 (4.22) | 65 (3.61) | 0.2062 |
| **Unsolicited adverse reactions within 0-7 days** | 52 (5.78) | 54 (5.99) | 106 (5.89) | 0.9203 |
| **Overall adverse reactions within 0-7 days** | 175 (19.47) | 167 (18.53) | 342 (19.00) | 0.6310 |

\* indicates that the P-value is calculated using Fisher's exact test. Data presents as n (%)

**Appendix Table A3:** Adverse reactions within 30 days of booster vaccination.

|  | NVSI-06-07<br>(N=899) | BBIBP-CorV<br>(N=901) | Total<br>(N=1800) | P value* |
| --- | --- | --- | --- | --- |
| **Solicited adverse reactions within 0-30 days** | 175 (19.47) | 160 (17.76) | 335 (18.61) | 0.3640 |
| Grade 1 | 152 (16.91) | 143 (15.87) | 295 (16.39) | 0.5670 |
| Grade 2 | 47 (5.23) | 31 (3.44) | 78 (4.33) | 0.0649 |
| Grade 3 | 1 (0.11) | 3 (0.33) | 4 (0.22) | 0.6246 |
| **Injection site adverse reactions** | 74 (8.23) | 50 (5.55) | 124 (6.89) | 0.0256 |
| Grade 1 | 65 (7.23) | 42 (4.66) | 107 (5.94) | 0.0220 |
| Grade 2 | 10 (1.11) | 8 (0.89) | 18 (1.00) | 0.6456 |
| Pain | 70 (7.79) | 47 (5.22) | 117 (6.50) | 0.0280 |
| Grade 1 | 61 (6.79) | 39 (4.33) | 100 (5.56) | 0.0237 |
| Grade 2 | 9 (1.00) | 8 (0.89) | 17 (0.94) | 0.8135 |
| Induration | 1 (0.11) | 0 | 1 (0.06) | 0.4994 |
| Grade 1 | 1 (0.11) | 0 | 1 (0.06) | 0.4994 |
| Swelling | 2 (0.22) | 1 (0.11) | 3 (0.17) | 0.6244 |
| Grade 1 | 2 (0.22) | 1 (0.11) | 3 (0.17) | 0.6244 |
| Erythra | 1 (0.11) | 2 (0.22) | 3 (0.17) | 1.0000 |
| Grade 1 | 1 (0.11) | 2 (0.22) | 3 (0.17) | 1.0000 |
| Redness | 1 (0.11) | 2 (0.22) | 3 (0.17) | 1.0000 |
| Grade 1 | 1 (0.11) | 2 (0.22) | 3 (0.17) | 1.0000 |
| Pruritus | 7 (0.78) | 3 (0.33) | 10 (0.56) | 0.2249 |
| Grade 1 | 6 (0.67) | 3 (0.33) | 9 (0.50) | 0.3421 |
| Grade 2 | 1 (0.11) | 0 | 1 (0.06) | 0.4994 |
| **Systemic adverse reactions** | 131 (14.57) | 130 (14.43) | 261 (14.50) | 0.9467 |
| Grade 1 | 109 (12.12) | 113 (12.54) | 222 (12.33) | 0.8298 |
| Grade 2 | 39 (4.34) | 26 (2.89) | 65 (3.61) | 0.1022 |
| Grade 3 | 1 (0.11) | 3 (0.33) | 4 (0.22) | 0.6246 |
| Fever | 23 (2.56) | 24 (2.66) | 47 (2.61) | 1.0000 |
| Grade 1 | 20 (2.22) | 21 (2.33) | 41 (2.28) | 1.0000 |
| Grade 2 | 2 (0.22) | 1 (0.11) | 3 (0.17) | 0.6244 |
| Grade 3 | 1 (0.11) | 3 (0.33) | 4 (0.22) | 0.6246 |
| Diarrhea | 3 (0.33) | 4 (0.44) | 7 (0.39) | 1.0000 |
| Grade 1 | 3 (0.33) | 4 (0.44) | 7 (0.39) | 1.0000 |
| Constipation | 4 (0.44) | 1 (0.11) | 5 (0.28) | 0.2177 |
| Grade 1 | 4 (0.44) | 1 (0.11) | 5 (0.28) | 0.2177 |
| Anorexia | 0 | 1 (0.11) | 1 (0.06) | 1.0000 |
| Grade 1 | 0 | 1 (0.11) | 1 (0.06) | 1.0000 |
| Vomiting | 2 (0.22) | 1 (0.11) | 3 (0.17) | 0.6244 |
| Grade 1 | 1 (0.11) | 1 (0.11) | 2 (0.11) | 1.0000 |
| Grade 2 | 1 (0.11) | 0 | 1 (0.06) | 0.4994 |
| Nausea | 4 (0.44) | 3 (0.33) | 7 (0.39) | 0.7258 |
| Grade 1 | 3 (0.33) | 3 (0.33) | 6 (0.33) | 1.0000 |
| Grade 2 | 1 (0.11) | 0 | 1 (0.06) | 0.4994 |
| Muscle pain (non-inoculation site) | 48 (5.34) | 41 (4.55) | 89 (4.94) | 0.4490 |
| Grade 1 | 38 (4.23) | 35 (3.88) | 73 (4.06) | 0.7218 |
| Grade 2 | 10 (1.11) | 7 (0.78) | 17 (0.94) | 0.4782 |
| Joint pain | 4 (0.44) | 3 (0.33) | 7 (0.39) | 0.7258 |
| Grade 1 | 3 (0.33) | 3 (0.33) | 6 (0.33) | 1.0000 |
| Grade 2 | 1 (0.11) | 0 | 1 (0.06) | 0.4994 |
| Headache | 49 (5.45) | 59 (6.55) | 108 (6.00) | 0.3718 |
| Grade 1 | 34 (3.78) | 45 (4.99) | 79 (4.39) | 0.2498 |
| Grade 2 | 16 (1.78) | 14 (1.55) | 30 (1.67) | 0.7176 |
| Cough | 14 (1.56) | 6 (0.67) | 20 (1.11) | 0.0765 |
| Grade 1 | 9 (1.00) | 4 (0.44) | 13 (0.72) | 0.1776 |
| Grade 2 | 6 (0.67) | 3 (0.33) | 9 (0.50) | 0.3421 |
| Breathing trouble | 3 (0.33) | 0 | 3 (0.17) | 0.1244 |
| Grade 1 | 3 (0.33) | 0 | 3 (0.17) | 0.1244 |
| Pruritus at non-inoculated sites (no skin damage) | 5 (0.56) | 5 (0.55) | 10 (0.56) | 1.0000 |
| Grade 1 | 3 (0.33) | 4 (0.44) | 7 (0.39) | 1.0000 |
| Grade 2 | 2 (0.22) | 1 (0.11) | 3 (0.17) | 0.6244 |
| Acute allergic reaction | 4 (0.44) | 2 (0.22) | 6 (0.33) | 0.4520 |
| Grade 1 | 2 (0.22) | 2 (0.22) | 4 (0.22) | 1.0000 |
| Grade 2 | 2 (0.22) | 0 | 2 (0.11) | 0.2493 |
| Fatigue | 30 (3.34) | 40 (4.44) | 70 (3.89) | 0.2725 |
| Grade 1 | 22 (2.45) | 37 (4.11) | 59 (3.28) | 0.0630 |
| Grade 2 | 8 (0.89) | 3 (0.33) | 11 (0.61) | 0.1444 |
| **Unsolicited adverse reactions within 0-30 days** | 57 (6.34) | 58 (6.44) | 115 (6.39) | 1.0000 |
| Grade 1 | 41 (4.56) | 46 (5.11) | 87 (4.83) | 0.6605 |
| Grade 2 | 18 (2.00) | 14 (1.55) | 32 (1.78) | 0.4825 |
| **Overall adverse reactions within 0-30 days** | 184 (20.47) | 177 (19.64) | 361 (20.06) | 0.6805 |
| Grade 1 | 162 (18.02) | 160 (17.76) | 322 (17.89) | 0.9022 |
| Grade 2 | 54 (6.01) | 40 (4.44) | 94 (5.22) | 0.1394 |
| Grade 3 | 1 (0.11) | 3 (0.33) | 4 (0.22) | 0.6246 |

\* indicates that the P-value is calculated using Fisher's exact test. Data presents as n (%)

## Notes

### Clinical Trial

NCT05033847

### Funding Statement

This study did not receive any funding

### Author Declarations

The trial protocol was reviewed and approved by Abu Dhabi Health Research and Technology Ethics Committee.

